# A mixed methods study on men’s and women’s tuberculosis care journeys in Lusaka, Zambia - implications for gender-tailored tuberculosis health promotion and case finding strategies

**DOI:** 10.1101/2022.11.17.22282462

**Authors:** Andrew D. Kerkhoff, Chanda Mwamba, Jake M. Pry, Mary Kagujje, Sarah Nyangu, Kondwelani Mateyo, Nsala Sanjase, Lophina Chilukutu, Katerina A. Christopoulos, Monde Muyoyeta, Anjali Sharma

## Abstract

**Background:** Men and women with undiagnosed tuberculosis (TB) in high burden countries may have differential factors influencing their healthcare seeking behaviors and access to TB services, which can result in delayed diagnoses and increase TB-related morbidity and mortality.

**Methods:** A convergent, parallel, mixed-methods study design was used to explore and evaluate TB care engagement among adults (≥18 years) with newly diagnosed, microbiologically-confirmed TB attending three public health facilities in Lusaka, Zambia. Quantitative structured surveys characterized the TB care pathway (time to initial care-seeking, diagnosis, and treatment initiation) and collected information on factors influencing care engagement. Multinomial multivariable logistic regression was used to determine predicted probabilities of TB health-seeking behaviors and determinants of care engagement. Qualitative in-depth interviews (IDIs; n=20) were conducted and analyzed using a hybrid approach to identify barriers and facilitators to TB care engagement by gender.

**Results:** Overall, 400 TB patients completed a structured survey, of which 275 (68.8%) and 125 (31.3%) were men and women, respectively. Men were more likely to be unmarried (39.3% and 27.2%), have a higher median daily income (50 and 30 Zambian Kwacha [ZMW]), alcohol use disorder (70.9% [AUDIT-C score ≥4] and 31.2% [AUDIT-C score ≥3]), and a history of smoking (63.3% and 8.8%), while women were more likely to be religious (96.8% and 70.8%) and HIV-positive (70.4% and 36.0%). After adjusting for potential confounders, the adjusted probability of delayed health-seeking ≥4 weeks after symptom onset did not differ significantly by gender (44.0% and 36.2%, p=0.14). While the top reasons for delayed healthcare-seeking were largely similar by gender, men were more likely to report initially perceiving their symptoms as not being serious (94.8% and 78.7%; p=0.032), while women were more likely to report not knowing the symptoms of TB before their diagnosis (89.5% and 74.4%; p=0.007) and having a prior bad healthcare experience (26.4% and 9.9%; p=0.036). Notably, women had a higher probability of receiving TB diagnosis ≥2 weeks after initial healthcare seeking (56.5% and 41.0%, p=0.007). While men and women reported similar acceptability of health-information sources, they emphasized different trusted messengers. Also, men had a higher adjusted probability of stating that no one influenced their health-related decision making (37.9% and 28.3%, p=0.001). IDIs largely corroborated the quantitative findings while offering more context and in-depth understanding of the factors that affected initial health seeking decisions, diagnoses, and treatment experiences across each step of men’s and women’s TB care pathways. To improve TB detection, men recommended TB testing sites at convenient community locations, while women endorsed an incentivized, peer-based, case-finding approach. Sensitization and TB testing strategies at bars and churches were highlighted as promising approaches to reach men and women, respectively.

**Conclusions:** Men and women with TB differ with respect to TB risk factors, TB care engagement experiences and determinants, and broader health influences. These differences suggest that gender-tailored TB health promotion and case-finding strategies may be needed to improve TB diagnosis and care engagement in high burden settings.

## Background

In 2020, more than 4 million persons with tuberculosis (TB) globally remained undiagnosed or went unreported,^1^ suggesting that a large proportion never received life-saving treatment. That same year, 56% of all TB notifications were among men, notwithstanding significantly higher gaps in case detection when compared to women.^1^ These data are consistent with a meta-analysis of TB prevalence surveys from 28 countries that found a 2.2-times higher prevalence of TB disease among men than in women.^2^ Further, men with TB may demonstrate lower rates of sputum conversion and cure,^3,4^ and are more likely to die.^5,6^ In some settings, gender disparities in TB-related outcomes, in part, appear to be underpinned by men’s late(r) presentation to TB services, as well as poorer retention in TB treatment services.^7–9^ Thus strategies to increase TB case detection require targeted approaches grounded in gendered differences in the TB epidemic and care pathway.

In order to design interventions and healthcare improvement strategies to better reach, appeal to, and overcome unique barriers faced by both men and women with undiagnosed TB, it is crucial to better understand the factors influencing TB care seeking and engagement in high burden settings, and whether they differ by gender. Across different settings, both men and women with undiagnosed TB may face substantial challenges seeking care for their illness and accessing TB diagnostic services.^5,8–10^ These may include insufficient TB-related knowledge (including its symptoms, where to go for services and the benefits of early diagnosis and treatment), insufficient time and/or money for transport and/or services (especially among the socioeconomically vulnerable), geographic inaccessibility, a lack of social support and self-efficacy, TB- (and HIV-) related stigma, privacy concerns, prior bad experiences with healthcare (including rude providers), or in the case of men, traditional views of masculinity that impede healthcare seeking. The determinants influencing engagement in TB care services in sub-Saharan Africa among men and women may be distinct, but to-date gender-specific differences across the TB care pathway, if present, are not well-characterized.

We undertook a mixed-methods study in Lusaka, Zambia, to explore and characterize care journeys for men and women with newly diagnosed TB. We sought to (a) evaluate TB risk factors, (b) understand barriers and facilitators to healthcare seeking after symptom onset, getting TB illness diagnosed, and starting TB therapy, and (c) to explore preferences for being reached with health information and potential strategies to improve TB detection. The primary objective of this study was to determine if and how these differ for men and women with TB. In the discussion section we use insights on similarities and differences to suggest gender-tailored strategies that may be needed to improve the key steps within the TB care pathway (**Fig 1)** and address any gender-specific inequities.

**Fig 1.**
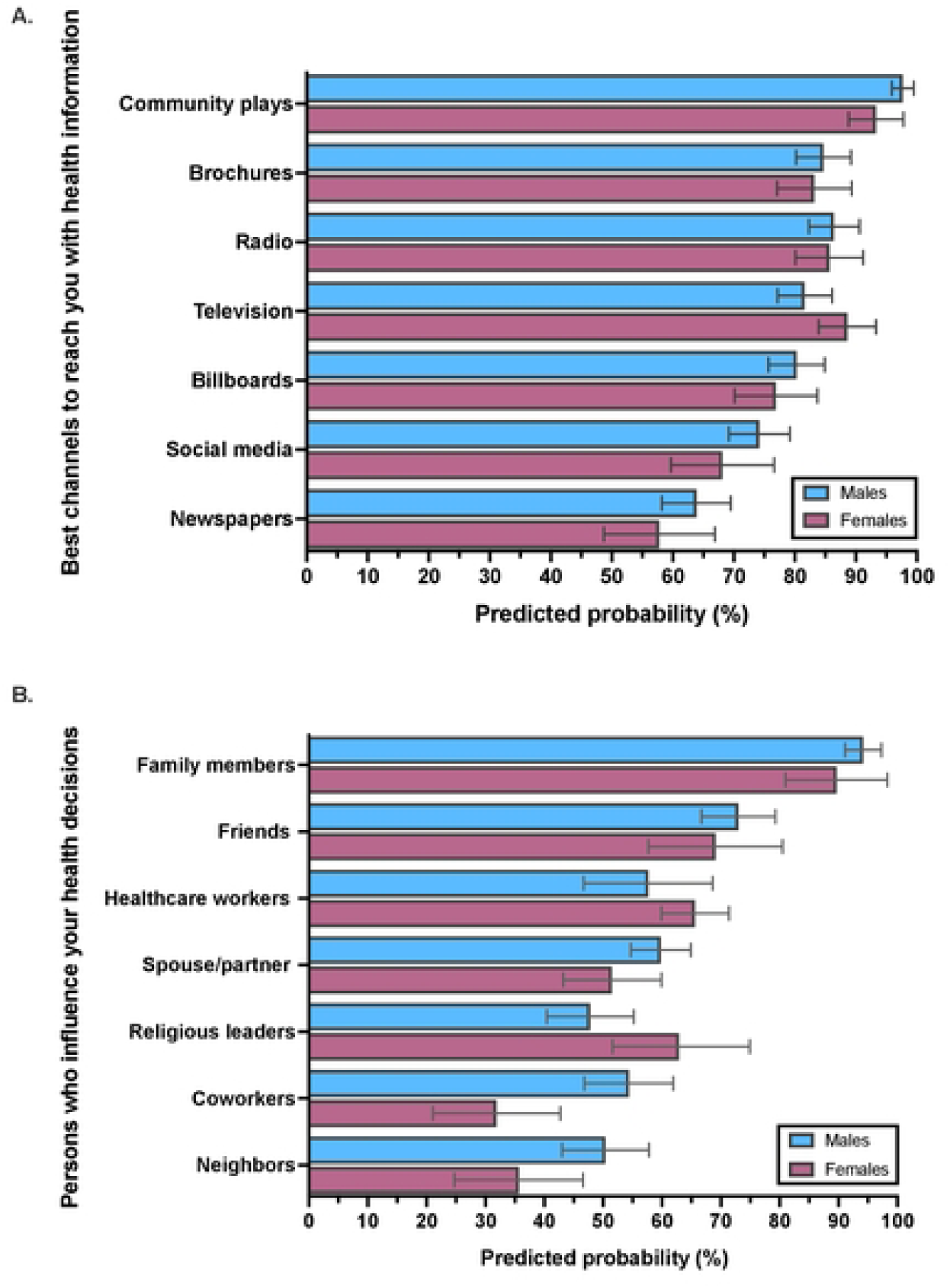
Conceptual framework for the mixed-methods study showing key steps in the care journey for persons with tuberculosis and superimposed key findings by gender from the combined analysis of quantitative and qualitative data.

## Methods

A convergent, parallel, mixed-methods design was used to yield a more complete understanding of complex experiences and multi-level barriers to care engagement among individuals with undiagnosed TB, by combining quantitative and qualitative data to construct complementary perspectives.^11,12^

### Setting and participants

The study was conducted at three public health facilities in Lusaka, Zambia from September 2019 to June 2020. Kanyama and Matero first-level hospitals have extremely busy outpatient TB clinics (1,263 and 1,522 TB notifications, respectively in 2021) and serve as the primary health facility for adjacent, densely populated, peri-urban townships; University Teaching Hospital (UTH), is a tertiary health facility serving Lusaka, and a referral hospital for Zambia. Quantitative data collection took place at Kanyama and University Teaching Hospitals, and qualitative data collection at all three facilities.

Persons with TB (i.e., TB patients) were eligible to participate in either quantitative or qualitative study activities if they were at least 18 years of age and had microbiologically-confirmed TB diagnosed in the last three weeks (to improve recall). An approximately equal number of participants were purposefully sampled from each of the three facilities, balanced for sex and HIV status. Recruitment for the quantitative data collection phase took place from September 18, 2019 to January 17, 2020, and closed once the pre-specified sample size was reached (∼n=400 participants); recruitment for the qualitative data collection phase took place from February 20^th^ 2020 to June 6^th^ 2020, and closed once data saturation was reached (10 men and 10 women for n=20).^12^ There were no individuals who participated in both the quantitative and qualitative data collection phases.

### Ethics, consent, and permissions

All participants provided written or verbal (for phone interviews) informed consent in their preferred language (Nyanja, Bemba or English). The University of Zambia Biomedical Research Ethics Committee (#010-014-19) and the institutional review board of the University of California, San Francisco (#18–26028) approved this study.

### Data collection

Both quantitative and qualitative data collection tools sought to explore and characterize men’s and women’s TB care journeys (**Fig 1)**.^13^ For the quantitative phase of the study, all participants completed a structured survey (**S1 Appendix)**. In addition to socio-demographic and clinical details, surveys captured information related to their pathway to TB care services, including timing of symptom onset, factors that may have influenced care-seeking and diagnosis, where and when they sought care, and their experience and satisfaction with TB services; whenever possible, validated measures for the survey were utilized.^14–16^ Surveys were administered via a touchscreen tablet using District Health Information Software 2 (DHIS2) with the help of a study team member. TB diagnosis and treatment details, as well as HIV status (reflecting either self-report or test result status if previously unknown) were extracted from routine, facility-specific, TB registers.

For the qualitative study phase, since recruitment took place around the start of the COVID-19 epidemic, the 20 TB patients completed in-depth interviews (IDIs) by phone to protect the safety of both the patients and study staff.^17^ A semi-structured guide, developed and structured according to key steps in TB care pathways,^18,19^ was used for the IDIs (**S2 Appendix)**. The guide used open-ended questions and incorporated probes based on the Capability, Opportunity, Motivation, Behavior (COM-B) theory of behavior change to clarify responses and encourage elaboration.^20^ Four experienced Zambian research assistants (RAs) collected the data after being trained in the study data collection methods. During training, the interview guide was practiced in the three interview languages (Nyanja, Bemba and English) to elicit participant experiences in the manner intended. During interviews, the interviewer captured the key points and developed the notes into a semi-structured memo immediately after the interview. Each IDI was audio-recorded and lasted 40-90 minutes.

### Data analysis and definitions

Alcohol use disorder was defined as an AUDIT-C score of ≥4 among men and ≥3 among women.^21^ For both quantitative and qualitative data analyses, we refer to gender-specific differences because we explored and characterized health-related behaviors and choices and the factors influencing those behaviors and choices (including social, cultural, and gender norms) among men and women with TB, rather than any differences that may be due to biology due to biology.^22^

### Quantitative analysis

For quantitative data analysis, because we were *a priori* interested in evaluating whether there were differences between men and women with TB, all analyses were undertaken overall and stratified by gender. Fisher’s exact, Pearson’s chi-squared, or Wilcoxon rank-sum tests were applied to compare socio-demographic characteristics among indicators with few responses, categorical covariates, and continuous covariates, respectively. Multinomial multivariable logistic regression was then used to determine the adjusted, predicted probabilities of specific TB health-seeking behaviors and determinants of care engagement for men and women. A directed acyclic graph (**S1 Figure**) was used to identify a minimally sufficient adjustment set of confounders and this informed the final multivariable model;^23^ in addition to the dependent TB healthcare-seeking determinant variable and the independent variable of interest, gender (male or female), models were adjusted for age, enrolment site, HIV status, education level, daily income, relationship status, alcohol use disorder status (AUDIT-C positive), history of prior TB, and knowing friends or family members who have had TB. Complete case analysis was conducted including only individuals for which complete survey data was recorded. All statistical tests were two-sided at alpha=0.05 and were conducted using Stata version 17.0 (StataCorp LLC, College Station, TX, USA).

### Qualitative analysis

Interviews were transcribed verbatim and directly translated into English. The RAs quality checked their own transcripts and CM quality checked all the transcripts. All the transcripts and memos were uploaded into Dedoose v7.0.23 (SocioCultural Research Consultants, Los Angeles, CA, USA) for coding. A hybrid process of inductive and deductive thematic analysis was followed.^24,25^ First, ADK created deductive codes from prior research evaluating gendered differences in TB care engagement.^5,9^ Next, CM, ADK and KC reviewed four transcripts and inductively developed codes (**S3 Appendix**). CM and ADK coded two transcripts each to refine the codes, followed by another two transcripts each to achieve consistency in coding. All transcripts were then coded by CM and ADK through an iterative process and categorized into themes. Analysis and interpretation of the data was categorized by gender to capture similarities and differences in perceptions, behaviors, experiences, and preferences.

### Triangulation

After analysis of the qualitative and quantitative data sets separately, ADK and CM compared the quantitative findings with the descriptive findings from the qualitative data for convergence. To relate and reflect on the gender-specific differences and suggested gender-tailored interventions, the quantitative statistical results were compared with the interpretation of each theme from the qualitative findings. The first authors reviewed and discussed the contradictions and the corroborated findings with related explanations.

## Results

### Overview of participants and characteristics by sex

Among 483 adults with confirmed TB approached by the study, 76 (15.7%) did not wish to participate (reasons were not collected), and 7 (1.4%) had incomplete survey data; therefore 400 (82.8%) were included in the quantitative data analysis. Overall, participants had a median age of 34 (IQR: 27-43) years, 275 (68.8%) were male, and 187 (46.8%) were HIV-positive (**Table 1**). The 20 TB patients who participated in the IDIs had a median age of 44 years (range, 23-67), 10 (50%) were male and female, respectively, and 10 (50%) were HIV-positive, 8 (40%) HIV-negative and 2 (10%) had an unknown HIV status.

**Table 1.** Characteristics of newly diagnosed tuberculosis patients in Lusaka, Zambia by sex (n=400)

|  | Overall<br>(n=400) | Male<br>(n=275) | Female<br>(n=125) | P-value |
| --- | --- | --- | --- | --- |
| **Site of enrolment** |  |  |  |  |
| First level hospital | 243 (60.8) | 191 (69.5) | 52 (41.6) | <0.001 |
| Tertiary hospital | 157 (39.3) | 83 (30.6) | 73 (58.4) |  |
| **Age in years, median (IQR)** | 34 (27-43) | 34 (27-42) | 34 (27-43) | 0.66 |
| **Age category** |  |  |  |  |
| 18-29.9 | 144 (36.0) | 102 (37.1) | 42 (33.6) | 0.79 |
| 30-39.9 | 141 (35.3) | 95 (34.6) | 46 (36.8) |  |
| ≥40 | 115 (28.8) | 78 (28.4) | 37 (29.6) |  |
| **Education** |  |  |  |  |
| None | 14 (3.5) | 8 (2.9) | 6 (4.8) | 0.70 |
| Primary | 148 (39.5) | 110 (40.0) | 48 (38.4) |  |
| Secondary | 192 (48.0) | 134 (48.7) | 58 (46.4) |  |
| Tertiary | 36 (9.0) | 23 (8.4) | 13 (10.4) |  |
| **Relationship status** |  |  |  |  |
| Currently married | 191 (47.8) | 124 (45.1) | 67 (53.6) | 0.005 |
| Divorced or separated | 49 (12.3) | 36 (13.1) | 13 (10.4) |  |
| Widowed | 18 (4.5) | 7 (2.6) | 11 (8.8) |  |
| Never married | 142 (35.5) | 108 (39.3) | 34 (27.2) |  |
| **Religious/faith** |  |  |  |  |
| Yes, regular church goer | 169 (42.3) | 75 (27.3) | 94 (75.2) | <0.001 |
| Yes, but not frequent church | 145 (36.3) | 118 (42.9) | 27 (21.6) |  |
| Not religious | 86 (21.5) | 82 (29.8) | 4 (3.2) |  |
| **Daily individual income (Kwacha), median (IQR)** | 50 (12-100) | 50 (30-100) | 30 (0-100) | <0.001 |
| **Primary income generator for household** |  |  |  |  |
| Yes | 266 (66.7) | 202 (73.5) | 64 (51.6) | <0.001 |
| No | 133 (33.3) | 73 (26.6) | 60 (48.4) |  |
| **Minibus use** |  |  |  |  |
| Every day | 71 (17.8) | 56 (20.4) | 15 (12.0) | 0.20 |
| 1-6 times per week | 100 (25.0) | 68 (25.1) | 31 (24.8) |  |
| A few times a month | 198 (49.5) | 129 (46.9) | 69 (55.2) |  |
| Never | 31 (7.8) | 21 (7.6) | 10 (8.0) |  |
| **HIV status** |  |  |  |  |
| Positive | 187 (46.8) | 99 (36.0) | 88 (70.4) | <0.001 |
| Negative | 213 (53.3) | 186 (64.0) | 37 (29.6) |  |
| **If HIV-positive, ART status** |  |  |  |  |
| Current daily use | 172 (92.5) | 90 (91.8) | 82 (93.2) | 0.47 |
| Current use, occasional missed doses | 5 (2.7) | 4 (4.1) | 1 (1.1) |  |
| Naïve | 9 (4.8) | 4 (4.1) | 5 (5.7) |  |
| **Ever tobacco smoker?** |  |  |  |  |
| Yes, daily | 151 (37.8) | 145 (52.7) | 6 (4.8) | <0.001 |
| Yes, less than daily | 34 (8.5) | 29 (10.6) | 5 (4.0) |  |
| No | 215 (53.8) | 101 (36.7) | 114 (91.2) |  |
| **AUDIT-C score (Alcohol Use), median (IQR)** | 4 (0-8) | 6 (2-10) | 0 (0-2) |  |
| **Alcohol use disorder (AUDIT-C positive)** |  |  |  |  |
| Yes | 234 (58.5) | 195 (70.9) | 39 (31.2) | <0.001 |
| No | 166 (41.5) | 80 (29.1) | 86 (68.8) |  |
| **How frequently do you go to a bar?** |  |  |  |  |
| Every day | 95 (23.8) | 80 (29.1) | 15 (12.0) | <0.001 |
| 1-6 times per week | 76 (19.0) | 70 (25.5) | 6 (4.8) |  |
| A few times a month | 98 (24.5) | 73 (26.6) | 25 (20.0) |  |
| Never | 131 (32.8) | 52 (18.9) | 79 (63.2) |  |
| ***TB history and contacts*** |  |  |  |  |
| **Past TB** |  |  |  |  |
| Yes | 56 (14.0) | 35 (12.7) | 21 (16.8) | 0.28 |
| No | 344 (86.0) | 240 (87.3) | 104 (83.2) |  |
| **Knows someone who has been diagnosed with TB** |  |  |  |  |
| Yes, last 6 months | 66 (16.5) | 49 (17.9) | 17 (13.6) | 0.016 |
| Yes, but not last 6 months | 123 (30.8) | 94 (34.3) | 29 (23.2) |  |
| No | 210 (52.6) | 131 (47.8) | 79 (63.2) |  |
| **Suspect TB in someone they know** |  |  |  |  |
| Yes, friend or family member | 106 (26.6) | 82 (29.9) | 24 (19.2) | 0.024 |
| No | 293 (73.4) | 192 (70.1) | 101 (80.8) |  |

The characteristics of survey participants stratified by gender did not differ by age, education, or prior history of TB (**Table 1)**. However, men with TB were more likely to report never being married (39.3% and 27.2%), higher median daily income (50 and 30 Zambian Kwacha (ZMW]) and being the primary household provider (73.5% and 51.6%). Conversely, women with TB were much more likely to report that they were religious (96.8% and 70.8%) and be HIV-positive (70.4% and 36.0%) than men with TB.

Men were also substantially more likely to have evidence of alcohol use disorder (i.e., AUDIT-C positive; 70.9% and 31.2%) and report a history of tobacco smoking (63.3% and 8.8%); of 234 participants with alcohol use disorder, 155 (79.5%) men and 8 (20.5%) women had a tobacco smoking history. Men were also much more likely to attend bars/taverns at least weekly (54.6% vs 16.8%) and 29.1% of men and 12.0% of women with TB reported going to a bar/tavern every day. Men were also more likely than women to say that they knew someone who had been previously diagnosed with TB (52.2% and 36.8%) and to suspect TB (not yet diagnosed) in a friend and/or family member (29.9% and 19.2%).

In the IDIs, men frequently attributed their TB disease to alcohol consumption and smoking ‘behavior’:

> P: “I had TB as a result of my behavior.”
>
> I: “If I may ask, what do you mean ‘as a result of’ what you were doing?
>
> P: “I was smoking.’’ (**Male, HIV negative**)

> ‘‘I don’t know how it (TB) started. I think it’s just the issue of drinking alcohol and smoking. I used to drink a lot and smoke whenever I went out.’’ (**Male, HIV positive**)

### TB health-seeking delays and care seeking experiences

#### Barriers to healthcare seeking

There was no gender-specific difference in the probability of delayed initial health-seeking ≥4 weeks after symptom onset (**Table 2;** 44.0% and 36.2%, p=0.14). Men and women were similarly likely to report that while they had considered getting help for their TB symptoms sooner, they consciously delayed healthcare seeking (53.6% and 54.6%, p=0.85). The top reasons provided for delayed care seeking were largely similar according to gender (**Fig 2a, S1 Table**). Both men and women commonly reported that they thought their symptoms were due to environmental causes such as pollution, that they preferred to try self-medication first, and that they lacked time to seek evaluation for their symptoms. However, men were more likely to report that they thought their symptoms were not serious initially and would spontaneously get better (94.8% and 78.7%, p=0.032), while women had a higher probability of stating that they did not know the symptoms of TB prior to their diagnosis (89.5% and 74.4%, p=0.007). While not a commonly cited reason for delayed health care seeking, women were also more likely than men to report prior bad healthcare experiences (26.4% and 9.9%, p=0.036).

**Table 2.** Tuberculosis care seeking behaviors and healthcare experiences among men and women with newly diagnosed tuberculosis in Lusaka, Zambia. Values represent adjusted predicted probabilities and associated 95% confidence intervals.

|  | **Overall**<br>**(95%CI)** | **Male**<br>**(95%CI)** | **Female**<br>**(95%CI)** | **Gender-specific**<br>**difference\***<br>**(95%CI)** |
| --- | --- | --- | --- | --- |
| **TB Health-Seeking** |  |  |  |  |
| **Health-seeking delay category**<br>**(Time from symptom onset to**<br>**first seeking care for illness)** |  |  |  |  |
| <4 weeks | 58.5 (54.1,63.0) | 56.0 (50.4,61.6) | 63.8 (55.5,72.0) | -7.8 (-18.3, 2.7) |
| 4-7.9 weeks | 30.6 (26.4,34.8) | 33.1 (27.6,38.6) | 25.7 (18.4,33.0) | 7.4 (-2.2, 17.1) |
| ≥8 weeks | 10.9 (7.8,13.9) | 10.9 (7.0,14.8) | 10.6 (5.0,16.2) | 0.3 (-6.9, 7.6) |
| **Location where participants**<br>**first visited for care related to**<br>**their illness** |  |  |  |  |
| Public clinic or hospital | 86.6 (83.4,89.7) | 88.0 (84.4,91.5) | 83.2 (74.3,92.0) | 4.8 (-5.0, 14.6) |
| Private clinic or hospital | 4.5 (2.4,6.5) | 3.5 (1.4,5.7) | 7.0 (1.9,12.1) | -3.5 (-9.3, 2.3) |
| Pharmacy | 7.8 (5.3,10.4) | 7.8 (4.9,10.7) | 8.0 (1.5,15.6) | -2.9 (-7.7, 7.1) |
| Herbalist/traditional healer | 1.0 (-) | 0.8 (-) | 1.8 (0,6.5) | -1.0 (-5.6, 3.7) |
| **TB Diagnosis** |  |  |  |  |
| **Diagnostic delay category (Time**<br>**from initial care-seeking to TB**<br>**diagnosis)** |  |  |  |  |
| <2 weeks | 54.4 (49.6,59.2) | 59.0 (53.3,64.8) | 43.5 (34.4,52.7) | 15.5 (4.1, 26.9) |
| 2-3.9 weeks | 25.4 (21.1,29.7) | 23.6 (18.4,28.7) | 28.7 (19.4,38.0) | -5.1 (-16.3, 6.1) |
| ≥4 weeks | 20.2 (16.4,24.0) | 17.4 (13.1,21.7) | 27.8 (18.5,37.1) | -10.4 (-21.7, 0.3) |
| **Diagnostics and treatment**<br>**received for TB illness prior to**<br>**diagnosis** |  |  |  |  |
| Antibiotics | 75.6 (20.3, 28.6) | 74.9 (69.6, 80.2) | 77.0 (69.2, 84.7) | -2.0 (-11.9, 7.9) |
| Blood test | 81.1 (77.6, 84.6) | 80.8 (76.6, 85.2) | 81.9 (76.4, 85.2) | 1.1 (-7.5, 9.6) |
| Chest x-ray | 74.3 (70.4, 78.2) | 74.3 (69.6, 79.0) | 74.4 (65.8, 83.0) | -0.2 (-10.4, 10.1) |
| Traditional remedy | 8.6 (5.9, 11.3) | 9.4 (5.8, 12.9) | 6.7 (1.9, 11.5) | 2.7 (-3.7, 9.0) |
| **Number of visits to**<br>**providers/facilities prior to**<br>**diagnosis** |  |  |  |  |
| 1 | 56.2 (51.3,61.0) | 58.1 (52.1,64.2) | 51.9 (42.2,61.5) | 6.2 (-5.8, 18.2) |
| 2 | 35.5 (30.9,40.2) | 33.8 (28.0,39.6) | 39.2 (29.8,48.6) | -5.4 (-17.1, 6.2) |
| ≥3 | 8.3 (5.7,11.0) | 8.1 (5.0,11.3) | 8.9 (3.0,14.8) | -0.8 (-7.8, 6.2) |
| **Time to diagnostic facility from**<br>**home** |  |  |  |  |
| <30 minutes | 40.1 (35.4,44.7) | 40.0 (34.2, 45.7) | 40.1 (40.7, 49.4) | -0.1 (-11.7, 11.4) |
| 30-60 minutes | 41.3 (36.6,46.0) | 43.3 (37.3, 49.3) | 37.4 (28.5, 46.3) | 6.0 (-5.4, 17.3) |
| >1-2 hours | 18.6 (14.9,22.4) | 16.7 (12.2, 21.3) | 22.5 (14.6, 30.5) | -5.8 (-15.5, 3.9) |
| **TB Treatment** |  |  |  |  |
| **Treatment delay category** |  |  |  |  |
| 0-2 days | 91.4 (88.8, 94.1) | 91.0 (87.7, 94.3) | 92.7 (87.5, 97.9) | -1.7 (-8.1, 4.7) |
| 3-5 days | 4.8 (2.7, 6.9) | 4.6 (2.1, 7.0) | 5.4 (0.8, 10.0) | -0.8 (-6.3, 4.6) |
| >5 days | 3.8 (2.0, 5.6) | 4.4 (2.1, 6.8) | 1.9 (0, 4.6) | 2.5 (-11.7, 6.2) |

\*Positive values indicate a higher probability among men, while negative values indicate a higher probability among women.

**Fig 2.**
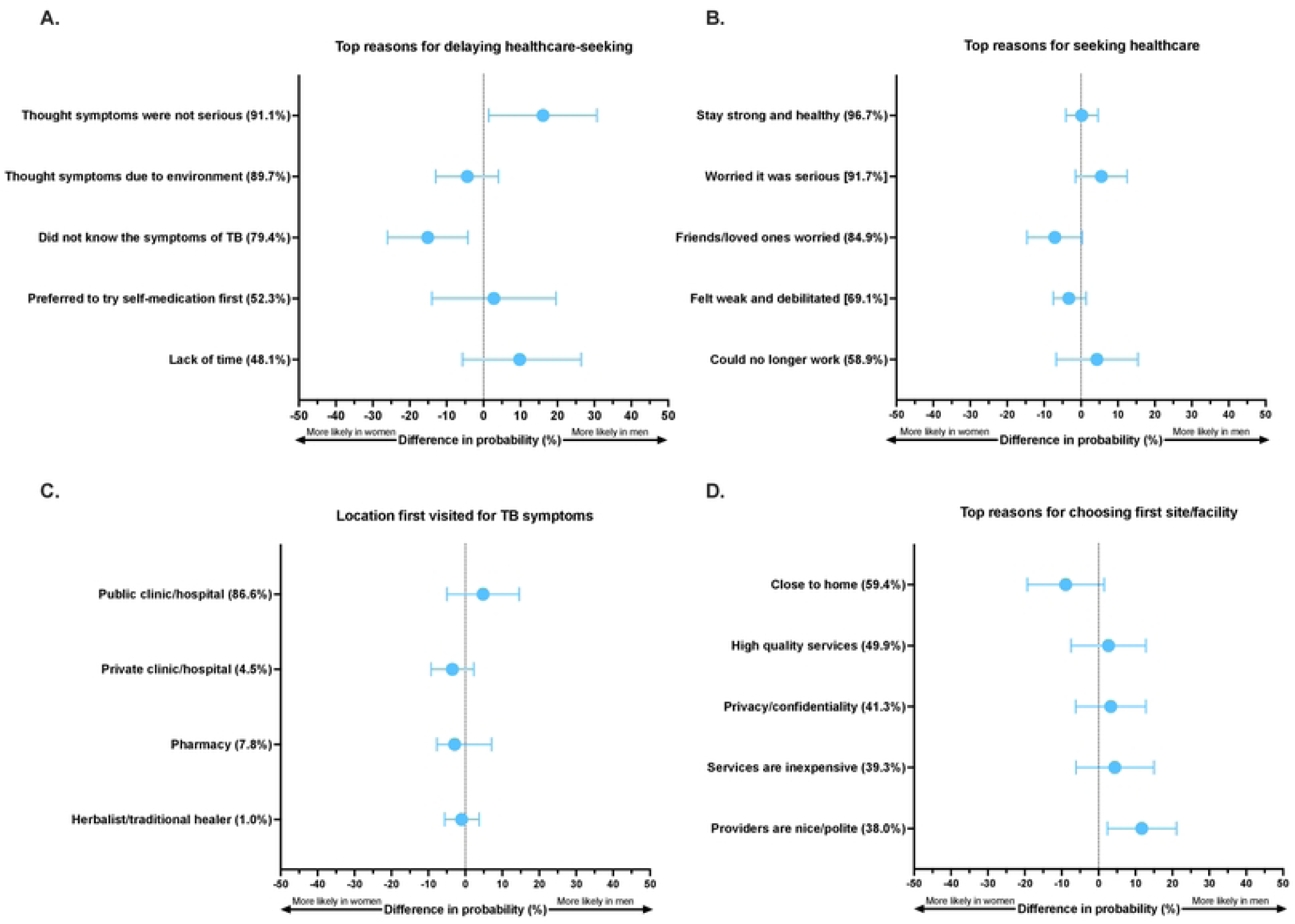
Gender specific differences in factors related to tuberculosis care engagement in Lusaka, Zambia among newly diagnosed tuberculosis patients: (a) Top reasons for delaying healthcare-seeking; (b) Top reasons for seeking healthcare for TB illness when they did; (c) Site or provider first visited for help with TB illness; (d) top reasons for choosing first site or provider. Positive values indicate factors are more probable among men, while negative values indicate factors are more probable among women.

Many IDI participants, irrespective of gender, corroborated the survey findings and reported “not worrying” about their cough and initially self-medicating with herbs and cough medicines based on advice from their peers, or their own initiative. They continued to take these medications, which temporarily alleviated some of their symptoms, and only visited the health facility when their symptoms did not improve, or they experienced additional symptoms, such as prolonged coughing fits, blood in their sputum, worsening chest pain, and excessive weight loss.

> ‘‘I didn’t even know what was happening. I was sweating after even having a bath. I tried drinking all sorts of medication for a cough, I thought I would get better and what misled me was the cough medicine. I was drinking [the medicine] but the cough was not finishing. That is when I decided to go to the clinic and they told me that my lungs were not fine and that is how I was put on TB medication’’ (**Female, HIV negative**)

Women, particularly those who were HIV-negative, disclosed that they lacked knowledge about TB symptoms and delayed seeking care because they ‘‘did not know what the problem was at that time and never thought about it [TB].’’

> I: “Before being found with TB, what did you know about TB?”
>
> P: “I really did not know about TB (…); when I was diagnosed with it, that is when I was taught about (TB).” (**Female, HIV negative**)

Men discussed delaying care seeking because of confusing their TB symptoms, such as a cough or weight loss, with the effects of excessive smoking and alcohol use. After onset of symptoms, they tried to first cut back on their smoking and alcohol intake. They only visited the health facility when symptoms persisted:

> ‘‘I used to smoke, and when I would cough, I would feel pain, and when I would talk about what I was feeling, I was told to stop smoking or else I would get TB’’ (**Male, HIV negative**)
>
> ‘‘So, people would say, ‘Ah big man! You have lost weight, are you not eating?’ Then I would say I have been drinking a lot of beer that is why. But when I reduced on beer, the same problem persisted, then I knew I had to go to the hospital.’’ (**Male, HIV negative**)

Some men who delayed seeking help cited conflicting work obligations as their reason. They believed that despite feeling unwell, men acted “fit” to keep their work commitments:

> ‘‘You just know how men behave; you keep on thinking you are fit and yet you are not okay. So, you’ll find that I would force myself to go for work even when I’m not feeling okay.’’ (**Male, HIV negative**)
>
> ‘‘For most men, one of the reasons [for delayed care seeking] is because they are the ones that work and take care of their families. Even if he is not feeling well and stays home, all eyes will be on him, because he needs to provide for his family. So even if he is not feeling well, he would rather go to work and keep on telling himself I’ll go to the clinic next time.’’ (Male, HIV positive)

Notably, some HIV-positive participants, regardless of gender, delayed seeking care because they thought their symptoms were related to HIV antiretroviral therapy (ART) side effects:

> I: “At that point that you lost weight, what did you think was happening to your body?”
>
> P: “I just thought the medicine [ART] was rejecting me (…), that is when I went to talk to my doctor. But it was not immediate, it took me almost a month.” (**Female, HIV positive**)

#### Factors influencing initial healthcare-seeking behaviors and location

Both men and women cited similar top reasons for ultimately seeking care for their TB illness (**Fig 2b, S1 Table**) including to stay healthy, being worried it was something serious, encouragement from worried loved ones and friends to get help, feeling debilitated, or no longer being able to work.

IDI participants similarly emphasized that they would visit a doctor if they had a serious illness. Both men and women indicated that symptoms, particularly a persistent cough with chest pain, “blood discharge” (i.e., hemoptysis), shortness of breath, and progressive weakness, led them to seek medical attention. Men were more likely to mention seeking treatment because they had lost too much weight, while women often mentioned having a persistent cough and being too weak to carry out their everyday tasks:

> “The only thing that made me go [to the clinic] was I was just noticing that I was lacking strength to do things and it was becoming so hard for me to even cut vegetables, and I would just sleep. I was failing even to fetch water in a bucket.” (**Female, HIV positive**)

Social support played a large role in influencing both men’s and women’s care seeking, with friends and families strongly encouraging them to seek help for their symptoms. Men were more likely to note that their spouses supported them by accompanying them to the health facility.

> ‘‘It’s the pain that I was having and the people who love me organized themselves and came to get me and I didn’t refuse. They came in and parked the vehicle because even when they were talking to me on phone my voice was very weak. So, we just went to the hospital so we could know the truth.’’ (**Male, HIV positive**)

Among men and women living with HIV, encouragement from their support network of peers, also living with HIV, was mentioned as a facilitator for seeking care:

> “There are some friends who are on medication (ART) and I even phoned them to ask if they also felt the same at some point and they encouraged me to visit the hospital because of how I was feeling.” (**Male, HIV positive**)

Notably, both male and female participants were highly likely to report they first presented to public health clinic or hospital for their TB illness (88.0% and 83.2%, p=0.33) **Fig 2c, Table 2**); additional sites of initial presentation (e.g., private health facilities, pharmacies, and traditional healers) were uncommon and did not differ by gender. Men and women noted similar factors that influenced the first site and/or provider they sought for help with their TB illness (**Fig 2d, S1 Table**); the most important reasons included proximity to home, perceived high quality services, privacy and confidentiality, inexpensive services, and because their friends/colleagues went there. Men were more likely than women to choose a facility because its providers were nice and polite (42.0 and 30.3%, p=0.014; **Fig 2d**).

Similarly, men and women in the IDIs often shared that they chose health facilities that were close to their homes to avoid transport costs and based on recommendations from friends and family members:

> I: “…Why did you go to xxx clinic?”
>
> P: “Because it is our nearest clinic and easiest to get there.” (**Female, HIV positive**)

On the other hand, some IDI participants said they avoided their local health facilities in favor of private clinics and higher-level government institutions to access specialized services, and to avoid poor perceived service, long queues, a purported lack of medical testing capacity and medication stockouts.

> “I wanted to go to xxx but people were saying the machines don’t work well they can’t even detect malaria. That is how I went to yyy and they tested me.” (**Male, HIV positive**)
>
> “I went there [private facility] because it is faster to get results there, at xxx queues are long. Just for you to be seen is a problem, and [t is] worse still if you are not feeling well. I wanted to be tested and told the results faster so that I could be told what I was suffering from to start medication.” (**Female, HIV negative**)

### TB diagnostic delays and experiences

After first seeking evaluation and care for the illness, women had a higher likelihood than men of experiencing more than 2-weeks of delay in being diagnosed with TB (56.5% and 41.0%, p=0.007, **Table 2**); notably 27.8% of women had diagnostic delays exceeding 4-weeks compared to 17.4% among men (p=0.06). There was no gender-specific difference in the likelihood of reporting visits to either 2 or 3-plus providers/facilities prior to their TB diagnosis being made (**Table 2)**. Men and women also had a similarly high probability of reporting that they received antibiotics and underwent blood tests, and chest radiography as part of their TB diagnostic journey (**Table 2**); use of traditional remedies was uncommon and didn’t differ by sex. There were few gender-specific differences in TB symptomatology (**S2 Table**), with only night sweats being slightly more likely among men (93.8% and 80.9%, p=0.007). Notably, the likelihood of reporting cough ≥2 weeks was similar between men and women (77.8% and 83.2, p=0.23), as was the probability of reporting ≥4 TB symptoms (92.5% and 91.6%, p=0.39).

In IDIs, both men and women corroborated findings of delayed TB diagnosis despite making multiple clinic visits. While they received cough medication, antibiotics, or prescriptions to purchase symptom-relieving medicines, they were tested for TB only when symptoms persisted. For example, a female participant claimed her TB remained undiagnosed for about two months while pregnant:

> **‘‘**I was pregnant and developed a really bad cough that I had for almost two months and that’s how I ended up giving birth to a premature baby because (…) it was difficult for them to find the TB. They would just give me Amoxil and say it’s just a cough and even when I finish the medication, it would not feel any better. I went to the hospital two times.’’ (**Female, HIV positive**)

Contrary to the survey, a few men and women disclosed initially seeking assistance from traditional healers, but that they found it unsatisfactory and later went to health facilities when they felt their symptoms were not improving:

> ‘‘Well at first, I tried the Traditional Healer but what he was saying was contrary to what I was feeling. So, I sat and thought that this problem could be sorted at the hospital and that is how I went to the hospital.’’ (**Male, HIV positive**)

### TB treatment delays and experiences

Overall, TB treatment delays were uncommon and did not differ by gender – where the probability of starting on TB therapy within 2 days of diagnosis (as confirmed by the TB register) was 91.0% and 92.7% for men and women, respectively (p=0.61; **Table 2)**.

In IDIs, both men and women corroborated that once diagnosed, they were quickly started on TB treatment. They reported that, in most cases, the results of TB tests were available the following day and that treatment was initiated the same or following day of TB diagnosis:

> I: “How long after being diagnosed did you start treatment?”
>
> P: “They gave me medication the same day I was diagnosed.” (**Female, HIV negative**)

IDI participants revealed that they had received information on the TB prevention, care, and treatment process after being diagnosed. Men said they had been told to give up smoking and drinking:

> ‘When I reached, they told me you will start taking drugs and I took some even there and they oriented me about TB, how its spread and how to prevent it from [spreading to] people. This was through a video.’ (**Male, HIV positive**)
>
> ‘‘I was told things like in the case of beer drinking I had to stop and also smoking had to stop.” (**Male, HIV positive**)

Both men and women expressed acceptance of the diagnosis and the need to take medication due to a desire to get well:

> ‘‘I just accepted [it] and moved on, and I make sure I take my drug. It’s life, there is nothing you can do.’’ (**Female, HIV positive**)

However, after receiving a TB diagnosis, some men and women disclosed experiencing ‘‘depression’’ and ‘fear’ of having TB, sometimes because they associated TB with HIV:

> ‘‘I was scared … I was so scared, I just wondered where I contracted it from since I frequently test for HIV.’’ (Female, HIV negative)
>
> ‘‘At the time [of TB diagnosis], I felt depressed and could think a lot. Then afterward, I realized the solution was just to follow instructions because you can’t keep thinking about where you got it from, or who gave it to you, because it is too late.’’ (**Male, HIV positive**)

In the survey, men and women had a similar probability of reporting that they were satisfied with the TB services they received (95.1% and 95.4%, p=0.93) and that the facility met their health-related needs (92.9% and 88.9%, p=0.29) (**S3 Table**). However, women were more slightly likely than man to say that they would recommend the TB facility to a friend or loved one (96.1% and 91.0%, p=0.045).

Among IDI participants, men and women corroborated the above and expressed a high degree of satisfaction with their experiences with receiving TB services. They perceived health workers as kind and helpful in guiding them through the clinic processes. As a result, both men and women stated that they intended to advise and encourage their friends and family to visit the medical facilities for treatment if they felt ill.

> ‘‘They just welcomed us very well; they don’t shout or insult, even when going to get medicine for TB, they welcome us very well.’’ (**Female, HIV negative**)

### Designing TB care engagement strategies for men and women

#### Preferred health channels and trusted sources of health information

Among survey respondents, most communication channels for health-related information were highly acceptable to both men and women with TB (**Fig 3a, S4 Table)**. The most popular mediums for reaching participants with health-related information included community-based plays, brochures, radio, television, and billboards – only television was more likely to be reported by women (88.6% and 81.6%, p=0.046). Notably, men had a higher probability than women of reporting that no one influenced or helped them make health-related decisions (37.9% and 28.3%, p=0.001). Among persons who reported that someone influenced health-related decisions, men and women had a similarly high likelihood of reporting family members, friends, health care workers and spouses as persons who helped them make health-related decisions (**Figure 3b, S4 Table)**. However, men had a higher probability of reporting that coworkers (54.4% and 31.9%, p=0.002) and neighbors (50.4% and 35.7%, p= 0.042) influenced their health-related decisions, while women had a higher probability of reporting that religious leaders (62.9% and 47.7%, p=0.044) influenced their health-related decisions (**Figure 3b, S4 Table)**.

**Fig 3.**
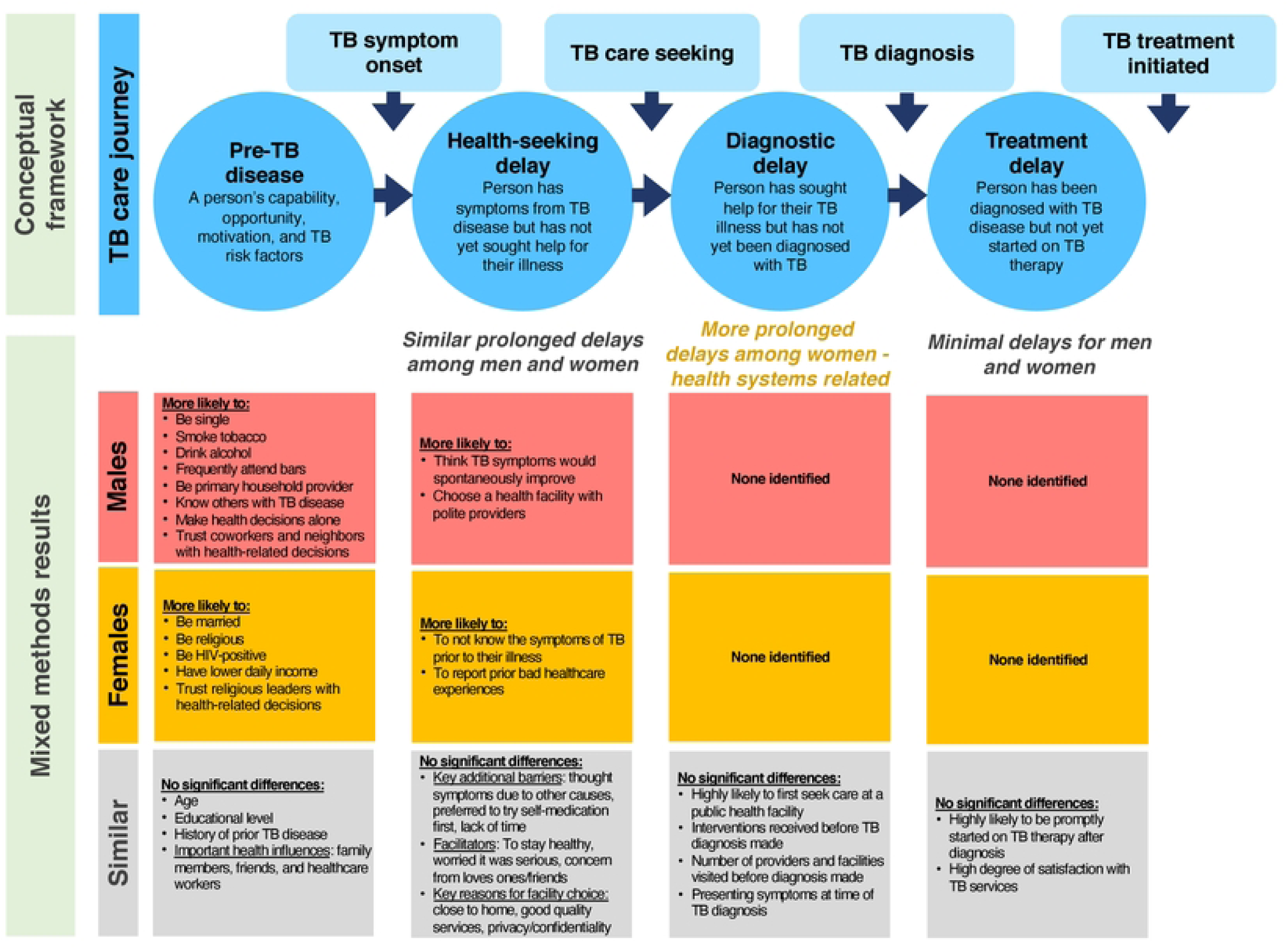
Preferred and trusted channels and messengers for health-related information among newly diagnosed tuberculosis patients according to gender: (a) Most preferred channels for being reached with health information; (b) Persons who influence their health-related decision making.

Both men and women in the IDIs also agreed that radio, television, SMS, and newspapers were effective tools for “educating” people about TB transmission and symptoms, as well as encouraging people to seek treatment for TB symptoms. In order to “reach” everyone who might not have access to radios and televisions, they also highlighted the importance of community-based engagement strategies:

> ‘‘It will be good for people to sensitize in the community because not everyone has a television and radio.’’ (**Male, HIV positive**)

#### Recommendations for improving TB detection among men and women

IDI participants were asked to consider three different potential strategies for improving TB detection in their community and state which one they thought might be most successful (**Table 3)**. About half of participants chose putting stands and mobile clinics offering TB testing in community places like markets and bus stations as the best strategy to improve TB detection; this was also the most preferred strategy among men, especially because of its convenience if one is simply “passing by” and is sick. Participants, especially women, also felt a promising strategy was to provide a small incentive to reach out to and encourage TB testing among peers that may have TB; this was because of their lived experience and ability to credibly teach others about TB. Few participants preferred a strategy that would provide persons with a small incentive to undergo TB testing at the local health facility.

**Table 3.** Perspectives and preferences of newly diagnosed tuberculosis patients for three potential strategies to improve tuberculosis diagnosis in their community (n= 17).

| Strategy and description | Number who stated that this strategy was most likely to be effective | Quotes |
| --- | --- | --- |
| **1. Place a mobile clinic or stand at a convenient location in the community (e.g., market, minibuss stand), to provide rapid TB testing** | Overall: 8<br>Men: 5<br>Women: 3 | "What is important is putting mobile clinics because they will be near people and when someone is just passing (by) they may think of testing for TB. Just like people go for cervical cancer, you will find that one is just coming from the market, but once they see the mobile clinic, they will turn and test, so it helps. (Male, HIV positive)<br>"The suggestion to open a mobile clinic is a good idea because some people will give reasons like not being able to go to the clinic to get tested because sometimes there are long distances to get to the clinic..." (Male, HIV negative)<br>"The best is that one of going into like the market or bus stops, because people are scared to go to the clinic, so you go and test them from there." (Female, HIV positive) |
| **2. Provide a small amount of money to get tested for TB at the health facility** | Overall: 2<br>Men: 1<br>Women: 1 | "Yes! ... You know people, where there is money, they can accept anything. You offer some money, packets of sugar, some drinks... you know this poverty in Zambia." (Male, HIV negative)<br>"It's good because some people become suffer(ing) from TB and (become) so sick that they have no means of looking for food." (Female, HIV positive) |
| **3. Provide brief training to current TB patients and give them a small amount of money to reach out to their peers that may have TB to encourage testing** | Overall: 7<br>Men: 2<br>Women: 5 | "If you give (training and incentives) to TB patients, they will also teach others who are seem(ing) to be sick. They will go and teach them and encourage them, tell them to go to the clinic and get tested... It is very much number one because they are able to encourage someone of their status, like what that friend of mine did to me, just because she went through it..." (Female, HIV positive)<br>"If we teach the current (TB patients) and give them something, they might be able to assist those people who are with the same disease." (Male, HIV positive)<br>"You know what it does to you, so you want also others to know fast what it does." (Female, HIV negative) |

Note: 2 men did not provide responses to this question as their interviews were ended early, and 1 woman did not provide clear opinion for which strategy she thought may be most effective

IDI participants also recommended door-to-door household testing campaigns in the community to identify more persons with TB.

> ‘Yes, I think like I said, the door-to-door campaign, it can work. For example, you have this program for polio whereby nurses go door-to-door, and also put-up points where they give vaccinations for polio and other medicine to prevent blindness. Even TB you can do the same.’’ (**Male, HIV negative**)

Participants further recommended that undertaking case finding in bars and taverns could help better reach and diagnose TB among men, as they spend a lot of time there:

> I: “What ways can be used to reach men for them to be diagnosed early?
>
> P: Okay men are found in the taverns, if you start going around the community taverns, that would help.” (**Male, HIV negative**)

Women on the other hand were said to be ‘into church’ and approaching them at churches would help in reaching them and detecting TB sooner:

> ‘‘Women are not a problem. They normally meet in groups at church on certain days, so we can do well to follow them on those days to sensitize them about TB.’’ (**Female, HIV positive**)

## Discussion

In this mixed-methods study among more than 400 newly diagnosed TB patients in Lusaka, Zambia, we identified several important gender-specific differences influencing TB diagnosis and care (**Fig 1)**. Of note, a larger proportion of men engaged in behaviors that increased their risk for developing TB disease (e.g., high levels of alcohol consumption and smoking), while women more often reported experiencing substantial delays in the diagnosis of their TB disease after initially seeking help for their symptoms. Furthermore, we identified gender-specific differences in factors affecting men’s and women’s healthcare seeking behaviors, trusted persons who influence their health, and preferences for improving TB detection in their community. These differences have important implications for the design of future strategies to improve TB diagnosis and care engagement for both men and women living in high TB burden settings.

Except for one study among multi-drug resistant TB patients from South Africa, the prevalence of alcohol use disorder among persons with TB in our study was much higher than prior prevalence estimates among TB patients in sub-Saharan Africa.^26^ Alcohol use has a dose-response relationship with TB, where larger consumption amounts - likely through several mechanisms^27^ – is associated with an increasing risk of both developing^28–30^ and dying from TB.^29,31^ Smoking is also an important risk factor for developing TB,^32,33^ and combined with alcohol use, may portend to an even higher TB risk through a synergistic relationship.^34^ In high TB burden settings, like Zambia, there is clearly an important need to address harmful alcohol use (found among 63% of the men) and smoking (among 80% of men with alcohol use disorder) as a strategy for preventing TB and improving treatment outcomes.^26,31,35^ One potentially promising strategy to better reach men could include outreach at bars, taverns, and betting halls, as also suggested by our interviewees, to provide alcohol use and smoking use education and support resources as part of a broader TB sensitization and control strategy; however, this and other strategies require further study to determine their acceptability, feasibility and potential effectiveness.

Prior studies have reported variable gender-specific differences in patient-related delays in healthcare seeking for TB symptoms.^5,7^ Among both men and women, lack of knowledge and disease recognition (e.g., cough) as well perceptions of disease causation (environment, curse) and severity led to self-medication, lifestyle changes and use of herbalists/traditional healers, which delayed care-seeking. These findings suggest an important need for ongoing community-based TB education and sensitization campaigns to reach men and women alike. Such campaigns should leverage regular opportunities for persons living with HIV engaged in care – emphasizing their increased risk of TB, its associated symptoms, and the need for timely evaluation even if they suspect their illness is due to other causes.

Our results provide important insights into the optimum ways to reach men and women with TB and other health related information. Motivations and nudges such as staying strong and healthy, concern and encouragement from loved ones, and the inability to continue working may be leveraged in future TB communication strategies to promote timely healthcare seeking among both men and women. Plays in the community, brochures/pamphlets, and the radio made for appealing edutainment to both men and women. Interpersonal communication should leverage the influence of co-workers (workplace) and neighbors by providing group education in the workplace and neighborhood for men, and that of religious leaders through church-based outreach approaches that include religious leaders as key stakeholders and trusted messengers for health initiatives targeted at women. Examples of such strategies in the TB literature are limited,^36,37^ but there has been demonstrated success associated with including religious leaders in campaigns to bolster COVID-19 vaccination coverage in different settings.^38–40^ It was notably that male participants were far more likely to state that no one influenced their healthcare decisions; understanding what TB-related messages are the most resonant and motivating, and what channels are most likely to reach men impervious to interpersonal influence requires preference research methods; best-worst scaling, are well suited for this purpose.^41^

In our study, men were similarly likely as women to cite lack of time as a factor contributing to delayed healthcare seeking, despite being the primary breadwinner for their household. Being the main income provider may be tied to masculine identity and is often cited as an important reason for prolonged delays in healthcare seeking for TB among men.^5,9,42,43^ Notably, few men and women reported cost as a barrier to healthcare seeking for TB despite reporting a lower average daily income. This suggests that unlike other settings, women’s financial dependency did not result in them demoting their healthcare-related needs and delaying access to TB diagnostic services.^9^ TB detection strategies that offer convenience and timeliness – such as community-based testing locations, after hours testing availability, or fast-track access points at facilities are likely to appeal to both men and women. In interviews, men preferred TB testing services at community locations such as minibus stands or at the market for their convenience while women preferred a peer-based strategy in which brief training and a small incentive were provided to recently diagnosed TB patients to reach out to and recommend that their family members, friends and other close contacts get tested. Testing in community locations should be further explored for patient-centeredness, convenience, and overcoming barriers to timely TB diagnosis, especially as more sensitive, rapid, point-of-care, and non-sputum-based diagnostics become available in the future.^44,45^ Several pilots and implementation research projects have shown that community testing, peer-led, and social-network based TB case finding strategies,^46– 51^ are promising approaches - although their epidemiologic impact may be limited.^52^ Nonetheless, interventional implementation studies and/or quasi-experimental studies are needed to understand how feasible and effective they and other community-based case-finding strategies may be as part of existing TB control programs.^53^

Public healthcare facilities provided the settings for health seeking behavior for both men and women based on proximity, quality of services, privacy and confidentiality, and (lack of) cost. The role of actors, i.e., providers, was important to men who valued polite and respectful interactions, which has been previously identified an important determinant for HIV care engagement and retention in Zambia.^54,55^ Emphasizing the convenience, quality, privacy of TB services and politeness of providers should be incorporated into future communication and quality improvement strategies to increase care engagement among both men and women.

Though found in many different high burden settings,^8,9,56,57^ including a prior study from Lusaka, Zambia,^58^ it is not immediately clear what underpinned the prolonged time to diagnosis among women with TB in our study. Both men and women presented with similar symptoms and frequently to a public health facility where TB diagnostic services (or specimen referral) are routinely available. While it is possible that TB education post-diagnosis led to over-reporting of symptoms, another possible explanation is low-risk perception for TB in women (especially those that are HIV negative) among healthcare workers, which led them to consider alternative etiologies even when women presented multiple times with classic TB symptoms.^7^ To further investigate this finding and explanatory causes, the use of standardized patient visits may be considered,^59^ that could then inform appropriate quality improvement efforts, including provider trainings. Further, the implementation of systematic TB testing for all symptomatic persons presenting for any reason to health facilities in high burden community (i.e., intensified case finding), could help overcome any gender-related clinical biases, if present.^60–62^

Strengths of our study were the inclusion of a large number of consecutively enrolled, microbiologically-confirmed TB patients from two different health facilities. Further, the integration of qualitative data from TB patients helped to provide greater insight and rich context for men’s and women’s converging and diverging experiences, and factors influencing TB care engagement. However, our study did have some limitations. While our study findings are likely representative of Lusaka province, which notifies more than 40% of annual TB cases in Zambia,^63^ some barriers, behaviors – which may be tied to local cultural norms – and healthcare experiences, are likely to be context-specific and thus may not be generalizable to other urban and peri-urban African settings. Additionally, our study was conducted directly prior to and at the very outset of the COVID-19 pandemic; it is not clear how this world altering event has and will continue to influence and modify men’s and women’s TB engagement behaviors and experiences. However, a prior study during the early COVID pandemic in Zambia appeared to demonstrate that men and women continued to seek care for TB illness despite substantial uncertainties and fear.^17^ Further, because we recruited persons shortly after diagnosis, we were unable to explore determinants of TB therapy adherence and retention in treatment services; while findings to date across settings are heterogenous, males may be more likely to non-adherent and have poor TB treatment outcomes.^5^ Finally, many variables we utilized were by self-report, including alcohol use, smoking, seeking help from traditional healers, and delays in initial healthcare seeking after TB symptom onset; given the potentially stigmatizing nature of these behaviors and social desirability bias, which may differ by gender, the true prevalence may have been underestimated.

## Conclusions

We identified several important and marked gender-related differences among men and women with TB in Zambia, including a high prevalence of alcohol use disorder and tobacco smoking among men, excess diagnostic delays among women, as well as differential determinants of healthcare-seeking, trusted messengers, and preferred strategies for improving TB detection. Collectively, this suggests that gender tailored strategies, acknowledging men’s and women’s unique challenges, needs, and preferences, may be needed to improve TB diagnosis and care engagement in high burden settings.

## Data Availability

Data files will be made available upon publication from the Dryad database.

https://doi.org/10.7272/Q6NV9GG2

## Author contributions

ADK, MM, and AS designed the study with input from CM and KAC. CM, SN, KM, NS, MK, LC, and MM helped with project administration and data collection. ADK was responsible for funding acquisition. ADK and CM was responsible for the database. ADK, CM, JMP, KAC, and AS, designed the analyses and ADK and CM performed the analyses. All authors commented on drafts and approved the final version of the article. ADK takes responsibility for the integrity of this study and decision to publish these data.

## Acknowledgments

We would like to acknowledge and thank the study staff, the staff at the individual health facilities, and the study participants who made this work possible.

## Data Availability

Data files will be made available upon publication from the Dryad database.

## Figure Legend

**S1 Fig. Directed acyclic graph (DAG) of factors that influence healthcare-seeking for tuberculosis**

## REFERENCES

1. World Health Organization. Global Tuberculosis Report 2021. World Health Organization, Geneva; 2021.

2. Horton KC, MacPherson P, Houben Rmgj, White RG, Corbett EL. Sex Differences in Tuberculosis Burden and Notifications in Low- and Middle-Income Countries: A Systematic Review and Meta-analysis. PLOS Med. 2016;13(9):e1002119. doi:10.1371/journal.pmed.1002119

3. Mlotshwa M, Abraham N, Beery M, et al. Risk factors for tuberculosis smear non-conversion in Eden district, Western Cape, South Africa, 2007–2013: a retrospective cohort study. Bmc Infect Dis. 2016;16(1):365. doi:10.1186/s12879-016-1712-y

4. Gomes NM de F, Bastos MC da M, Marins RM, et al. Differences between Risk Factors Associated with Tuberculosis Treatment Abandonment and Mortality. Pulmonary Medicine. 2015;2015(6):1–8. doi:10.1155/2015/546106

5. Hoff SV den, Najilis CA, Bloss E, Straetemans M. A Systematic Review on the Role of Gender in Tuberculosis Control. KNCV Tuberculosis Foundation. 2010.

6. Imperial MZ, Nahid P, Phillips PPJ, et al. A patient-level pooled analysis of treatment-shortening regimens for drug-susceptible pulmonary tuberculosis. Nat Med. 2018;24(11):1708–1715. doi:10.1038/s41591-018-0224-2

7. Storla DG, Yimer S, Bjune GA. A systematic review of delay in the diagnosis and treatment of tuberculosis. BMC Public Health. 2008;8(1):15. doi:10.1186/1471-2458-8-15

8. Yang WT, Gounder CR, Akande T, et al. Barriers and Delays in Tuberculosis Diagnosis and Treatment Services: Does Gender Matter? Tuberculosis Research and Treatment. 2014;2014(3):1–15. doi:10.1155/2014/461935

9. Krishnan L, Akande T, Shankar AV, et al. Gender-Related Barriers and Delays in Accessing Tuberculosis Diagnostic and Treatment Services: A Systematic Review of Qualitative Studies. Tuberc Res Treat. 2014;2014(2):1–14. doi:10.1155/2014/215059

10. Muula AS. Gender differences in delays to TB diagnosis. The international journal of tuberculosis and lung disease : the official journal of the International Union against Tuberculosis and Lung Disease. 2001;5(11):1072–1074.

11. Dawadi S, Shrestha S, Giri RA. Mixed-Methods Research: A Discussion on its Types, Challenges, and Criticisms. J Pract Stud Educ. 2021;2(2):25–36. doi:10.46809/jpse.v2i2.20

12. Tariq S, Woodman J. Using mixed methods in health research. Jrsm Short Reports. 2013;4(6):2042533313479197. doi:10.1177/2042533313479197

13. Stop TB Partnership. TB Journeys: Our stories, our words. Stop TB, Geneva, 2016. Published online October 1, 2016.

14. Organization WH. Tuberculosis Patient Cost Surveys: a Handbook. World Health Organization, Geneva; 2017. http://apps.who.int/iris/bitstream/handle/10665/259701/9789241513524-eng.pdf?sequence=1

15. CDC. Tobacco Questions for Surveys - A Subset of Key Questions from the Global Adult Tobacco Survey (GATS) 2nd Edition. CDC, Atlanta; 2011.

16. Bush K, Kivlahan DR, McDonell MB, Fihn SD, Bradley KA. The AUDIT Alcohol Consumption Questions (AUDIT-C): An Effective Brief Screening Test for Problem Drinking. Arch Intern Med. 1998;158(16):1789–1795. doi:10.1001/archinte.158.16.1789

17. Mwamba C, Kerkhoff AD, Kagujje M, Lungu P, Muyoyeta M, Sharma A. Diagnosed with TB in the era of COVID-19: patient perspectives in Zambia. Public Heal Action. 2020;10(4):141–146. doi:10.5588/pha.20.0053

18. WHO. Diagnostic and treatment delay in tuberculosis - An in-depth analysis of the health-seeking behaviour of patients and health system response in seven countries of the Eastern Mediterranean Region. World Health Organization, Geneva; 2006.

19. Sreeramareddy CT, Qin ZZ, Satyanarayana S, Subbaraman R, Pai M. Delays in diagnosis and treatment of pulmonary tuberculosis in India: a systematic review. Int J Tuberc Lung Dis. 2014;18(3):255–266. doi:10.5588/ijtld.13.0585

20. Michie S, Stralen MM van, West R. The behaviour change wheel: A new method for characterising and designing behaviour change interventions. Implement Sci. 2011;6(1):42. doi:10.1186/1748-5908-6-42

21. Bradley KA, DeBenedetti AF, Volk RJ, Williams EC, Frank D, Kivlahan DR. AUDIT‐C as a Brief Screen for Alcohol Misuse in Primary Care. Alcohol Clin Exp Res. 2007;31(7):1208–1217. doi:10.1111/j.1530-0277.2007.00403.x

22. Nhamoyebonde S, Leslie A. Biological Differences Between the Sexes and Susceptibility to Tuberculosis. The Journal of Infectious Diseases. 2014;209(suppl 3):S100–S106. doi:10.1093/infdis/jiu147

23. Shrier I, Platt RW. Reducing bias through directed acyclic graphs. BMC Medical Research Methodology. 2008;8(1):7–15. doi:10.1186/1471-2288-8-70

24. Bradley EH, Curry LA, Devers KJ. Qualitative Data Analysis for Health Services Research: Developing Taxonomy, Themes, and Theory. Health Serv Res. 2007;42(4):1758–1772. doi:10.1111/j.1475-6773.2006.00684.x

25. Attride-Stirling J. Thematic networks: an analytic tool for qualitative research. Qual Res. 2001;1(3):385–405. doi:10.1177/146879410100100307

26. Necho M, Tsehay M, Seid M, et al. Prevalence and associated factors for alcohol use disorder among tuberculosis patients: a systematic review and meta-analysis study. Subst Abus Treat Prev Policy. 2021;16(1):2. doi:10.1186/s13011-020-00335-w

27. Wigger GW, Bouton TC, Jacobson KR, Auld SC, Yeligar SM, Staitieh BS. The Impact of Alcohol Use Disorder on Tuberculosis: A Review of the Epidemiology and Potential Immunologic Mechanisms. Front Immunol. 2022;13:864817. doi:10.3389/fimmu.2022.864817

28. Simou E, Britton J, Leonardi-Bee J. Alcohol consumption and risk of tuberculosis: a systematic review and meta-analysis. Int J Tuberc Lung Dis. 2018;22(11):1277–1285. doi:10.5588/ijtld.18.0092

29. Imtiaz S, Shield KD, Roerecke M, Samokhvalov AV, Lönnroth K, Rehm J. Alcohol consumption as a risk factor for tuberculosis: meta-analyses and burden of disease. Eur Respir J. 2017;50(1):1700216. doi:10.1183/13993003.00216-2017

30. Bates MN, Khalakdina A, Pai M, Chang L, Lessa F, Smith KR. Risk of Tuberculosis From Exposure to Tobacco Smoke: A Systematic Review and Meta-analysis. Arch Intern Med. 2007;167(4):335–342. doi:10.1001/archinte.167.4.335

31. Ragan EJ, Kleinman MB, Sweigart B, et al. The impact of alcohol use on tuberculosis treatment outcomes: a systematic review and meta-analysis. Int J Tuberc Lung Dis. 2020;24(1):73–82. doi:10.5588/ijtld.19.0080

32. Padrão E, Oliveira O, Felgueiras Ó, Gaio AR, Duarte R. Tuberculosis and tobacco: is there any epidemiological association? Eur Respir J. 2018;51(1):1702121. doi:10.1183/13993003.02121-2017

33. Murrison LB, Martinson N, Moloney RM, et al. Tobacco Smoking and Tuberculosis among Men Living with HIV in Johannesburg, South Africa: A Case-Control Study. Plos One. 2016;11(11):e0167133. doi:10.1371/journal.pone.0167133

34. Soh AZ, Chee CBE, Wang YT, Yuan JM, Koh WP. Alcohol drinking and cigarette smoking in relation to risk of active tuberculosis: prospective cohort study. Bmj Open Respir Res. 2017;4(1):e000247. doi:10.1136/bmjresp-2017-000247

35. Raviglione M, Poznyak V. Targeting harmful use of alcohol for prevention and treatment of tuberculosis: a call for action. Eur Respir J. 2017;50(1):1700946. doi:10.1183/13993003.00946-2017

36. Islam Z, Sanin KI, Ahmed T. Improving case detection of tuberculosis among children in Bangladesh: lessons learned through an implementation research. Bmc Public Health. 2017;17(1):131. doi:10.1186/s12889-017-4062-9

37. TB Europe Coalition. Engaging religious leaders in TB advocacy: experience of the initiative for health Foundation in Sofia, 2019.

38. Abdul-Mutakabbir JC, Casey S, Jews V, et al. A three-tiered approach to address barriers to COVID-19 vaccine delivery in the Black community. Lancet Global Heal. 2021;9(6):e749–e750. doi:10.1016/s2214-109x(21)00099-1

39. Johns Hopkins Center for Communications Channels. Engaging Religious Leaders to Boost COVID-19 Vaccination. 2022.

40. Privor-Dumm L, King T. Community-based Strategies to Engage Pastors Can Help Address Vaccine Hesitancy and Health Disparities in Black Communities. J Health Commun. 2021;25(10):827–830. doi:10.1080/10810730.2021.1873463

41. Soekhai V, Whichello C, Levitan B, et al. Methods for exploring and eliciting patient preferences in the medical product lifecycle: a literature review. Drug Discovery Today. 2019;24(7):1324–1331. doi:10.1016/j.drudis.2019.05.001

42. Chikovore J, Hart G, Kumwenda M, Chipungu GA, Corbett L. ‘For a mere cough, men must just chew Conjex, gain strength, and continue working’: the provider construction and tuberculosis care-seeking implications in Blantyre, Malawi. Global Health Action. 2015;8(1):26292. doi:10.3402/gha.v8.26292

43. Chikovore J, Pai M, Horton KC, et al. Missing men with tuberculosis: the need to address structural influences and implement targeted and multidimensional interventions. Bmj Global Heal. 2020;5(5):e002255. doi:10.1136/bmjgh-2019-002255

44. WHO. High-priority target product profiles for new tuberculosis diagnostics: report of a consensus meeting. World Health Organization, Geneva; 2014.

45. Drain PK, Gardiner J, Hannah H, et al. Guidance for Studies Evaluating the Accuracy of Biomarker-Based Nonsputum Tests to Diagnose Tuberculosis. J Infect Dis. 2019;220(Supplement_3):S108–S115. doi:10.1093/infdis/jiz356

46. Tuot S, Teo AKJ, Cazabon D, et al. Acceptability of active case finding with a seed-and-recruit model to improve tuberculosis case detection and linkage to treatment in Cambodia: A qualitative study. Galea JT, ed. PLOS One. 2019;14(7):e0210919. doi:10.1371/journal.pone.0210919

47. André E, Rusumba O, Evans CA, et al. Patient-led active tuberculosis case-finding in the Democratic Republic of the Congo. Bull World Health Organ. 2018;96(8):522–530. doi:10.2471/blt.17.203968

48. Joshi D, Sthapit R, Brouwer M. Peer-led active tuberculosis case-finding among people living with HIV: lessons from Nepal. Bull World Health Organ. 2017;95(2):135–139. doi:10.2471/blt.16.179119

49. McDowell M, Hossain M, Rahman N, et al. Expanding tuberculosis case notification among marginalized groups in Bangladesh through peer sputum collection. Public Health Action. 2015;5(2):119–121. doi:10.5588/pha.15.0014

50. Goldberg J, Macis M, Chintagunta P. Leveraging Patients’ Social Networks to Overcome Tuberculosis Underdetection: A Field Experiment in India. IZA Discussion Paper No 11942. Published online 2018.

51. Vo LNQ, Forse RJ, Codlin AJ, et al. A comparative impact evaluation of two human resource models for community-based active tuberculosis case finding in Ho Chi Minh City, Viet Nam. Bmc Public Health. 2020;20(1):934. doi:10.1186/s12889-020-09042-4

52. McCreesh N, White RG. An explanation for the low proportion of tuberculosis that results from transmission between household and known social contacts. Sci Rep. 2018;8(1):5382. doi:10.1038/s41598-018-23797-2

53. Burke RM, Nliwasa M, Feasey HRA, et al. Community-based active case-finding interventions for tuberculosis: a systematic review. Lancet Public Heal. 2021;6(5):e283–e299. doi:10.1016/s2468-2667(21)00033-5

54. Zanolini A, Sikombe K, Sikazwe I, et al. Understanding preferences for HIV care and treatment in Zambia: Evidence from a discrete choice experiment among patients who have been lost to follow-up. PLOS Med. 2018;15(8):e1002636. doi:10.1371/journal.pmed.1002636

55. Mwamba C, Sharma A, Mukamba N, et al. ‘They care rudely!’: resourcing and relational health system factors that influence retention in care for people living with HIV in Zambia. BMJ Glob Health. 2018;3(5):e001007. doi:10.1136/bmjgh-2018-001007

56. Gosoniu GD, Ganapathy S, Kemp J, et al. Gender and socio-cultural determinants of delay to diagnosis of TB in Bangladesh, India and Malawi. The international journal of tuberculosis and lung disease : the official journal of the International Union against Tuberculosis and Lung Disease. 2008;12(7):848–855.

57. Weiss MG, Sommerfeld J, Uplekar MW. Social and cultural dimensions of gender and tuberculosis. Int J Tuberc Lung Dis. 2008;12(7):829–830.

58. Needham DM, Foster SD, Tomlinson G, Godfrey-Faussett P. Socio-economic, gender and health services factors affecting diagnostic delay for tuberculosis patients in urban Zambia. Tropical Medicine & International Health. 2001;6(4):256–259.

59. Daniels B, Kwan A, Satyanarayana S, et al. Use of standardised patients to assess gender differences in quality of tuberculosis care in urban India: a two-city, cross-sectional study. Lancet Global Heal. 2019;7(5):e633–e643. doi:10.1016/s2214-109x(19)30031-2

60. Kagujje M, Chilikutu L, Somwe P, et al. Active TB case finding in a high burden setting; comparison of community and facility-based strategies in Lusaka, Zambia. PLoS ONE. Published online 2020.

61. Hanrahan CF, Nonyane BAS, Mmolawa L, et al. Contact tracing versus facility-based screening for active TB case finding in rural South Africa: A pragmatic cluster-randomized trial (Kharitode TB). PLOS Med. 2019;16(4):e1002796. doi:10.1371/journal.pmed.1002796

62. Stop TB. Intensified TB Case Finding at Facility Level. Stop TB Partnership, Geneva; 2018.

63. Kapata N, Chanda-Kapata P, Ngosa W, et al. The Prevalence of Tuberculosis in Zambia: Results from the First National TB Prevalence Survey, 2013–2014. Deribe K, ed. PLoS ONE. 2016;11(1):e0146392–14. doi:10.1371/journal.pone.0146392

